# Multi-Ancestry Genome-wide Association Analyses Identify Shared and Specific Genetic Architecture in Mild and Moderate-to-Severe Asthma

**DOI:** 10.64898/2026.09.20.26363488

**Authors:** Angelico Mendy, Yadu Gautam, Bradley H. Rosen, Joseph Castlen, Michael B. Fessler, Darryl C. Zeldin, Peter S. Thorne, Tesfaye B. Mersha

## Abstract

**Background:** Moderate-to-severe asthma affects 15-30% of asthma patients, but accounts for over 50% of healthcare costs and disproportionate morbidity. However, the genetic mechanisms underlying moderate-to-severe versus mild asthma remain poorly understood.

**Methods:** We performed separate pooled multi-ancestry GWAS of mild (N = 14,372) and moderate-to-severe asthma (N = 7,096) versus non-asthma controls (N = 28,816) among adult participants in the NIH All of Us Research Program (version 8). European, African, and Latino/admixed ancestry-specific GWAS were combined using fixed-effect meta-analysis. Lung expression quantitative trait loci (eQTL) and pathway analyses were performed to identify candidate genes and biological pathways across asthma phenotypes.

**Results:** Mild and moderate-to-severe asthma shared susceptibility loci at *IL1RL1*, *IKZF3*, and the *GTF3AP1-IL33*, *GTF3AP1-RANBP6*, *LINC02757-EMSY* and *HLA-DRB1-HLA-DQA1* regions. Shared lung eQTLs implicated *IL18R1*, *IL18RAP*, HLA genes, *IL33*, *LRRC32*, *GRB7*, *MIEN1* and *GSDMB*. In contrast, the phenotypes exhibited distinct genetic architectures. Mild asthma was characterized by broader HLA class II signals and associations at *SMAD3* and *GSDMB*, with enrichment of antigen presentation, T-helper cell differentiation, and TGF-β regulation, consistent with adaptive immune mechanisms. Moderate-to-severe asthma was characterized by *WDR36* and *PTCH1*, with lung eQTLs implicating *TSLP*, *CAMK4*, and *FANCC*. Pathway analysis identified enrichment of IL-13, IL-6, and IL-10 production, myeloid leukocyte differentiation, and oxidative stress responses, consistent with innate inflammation and pathways implicated in steroid resistance. The *IL33* signals showed a severity-gradient effect, with stronger associations in moderate-to-severe asthma.

**Conclusion:** Mild and moderate-to-severe asthma exhibit shared and distinct genetic architectures that support existing biologic targets and suggest novel therapeutic candidates.

## INTRODUCTION

Asthma is a chronic respiratory disease affecting 300 million people and causing more than 450,000 annual deaths worldwide.^1^ It is characterized by airway inflammation and bronchial hyperresponsiveness resulting in reversible airflow obstruction and airway remodeling that cause wheezing, shortness of breath, cough and chest tightness.^2^ Asthma severity ranges from mild to severe disease.^2^ Mild asthma is characterized by infrequent symptoms, preserved lung function and minimal treatment requirements, whereas moderate-to-severe asthma is associated with persistent symptoms requiring daily inhaled corticosteroids (ICS) combined with additional controller medications, biologic therapies or oral corticosteroids to achieve disease control.^2^ Moderate-to-severe asthma accounts for 15-30% of asthma cases but contributes disproportionately to healthcare burden and costs in the U.S.^3^ These patients are at increased risk of severe exacerbations and treatment-related adverse effects. High-dose ICS are associated with an increased risk of pneumonia and cardiovascular complications, while chronic oral corticosteroid use is associated with higher risk of osteoporosis, skin thinning, infections, cataracts, and cardiovascular, metabolic and renal complications.^4,5^

Both environmental and genetic factors contribute to asthma susceptibility, with heritability estimated at 35%-95%; however, most genome-wide association studies (GWAS) have focused on asthma case-control status rather than disease severity.^6^ Only four GWAS have specifically investigated moderate-to-severe asthma despite its substantial clinical burden, including two with fewer than 1,000 cases.^7–10^ The TENOR study (473 cases) identified associations at *RAD50-IL13* and *HLA-DR/DQ*.^7^ The AUGOSA study (933 cases) replicated the *ORMDL3/GSDMB* locus.^8^ A recent GWAS of 3,181 cases identified a lung function locus near *THSD4*.^9^ The largest GWAS to date (5,135 discovery and 5,414 replication cases) identified *MUC5AC* as a severity-specific locus and concluded that mild and moderate-to-severe asthma share substantial genetic architecture based on comparisons with previous asthma studies.^10^ However, all previous moderate-to-severe asthma GWAS were limited to European ancestry populations and did not directly compare the genetic architectures of mild and moderate-to-severe asthma. We therefore conducted the first multi-ancestry GWAS of moderate-to-severe asthma alongside a parallel GWAS of mild asthma to identify shared and severity-specific genetic risk factors.

## METHODS

### Data Source

We used data from the National institutes of Health (NIH) All of Us Research Program (AoURP) data version 8, a prospective cohort recruiting ≥1 million U.S. participants aged ≥18 years to investigate the effects of lifestyle, environmental and genetic factors on health.^11^ AoURP prioritizes populations underrepresented in biomedical research, including racial, ethnic, sexual and gender minorities, low-income people, rural residents.^11^ Recruitment began in May 2018 through healthcare provider organizations and direct volunteer enrollment.^11^ Participants provided informed consent, completed baseline surveys, authorized electronic health record (EHRs) access and optionally contributed biospecimens for genomic analyses.^11^ As of June 2026, 633,248 participants have enrolled.

### Mild and Moderate-to-severe Asthma

Asthma was identified from EHRs using Systematized Nomenclature of Medicine (SNOMED) and International Classification of Diseases, Tenth Revision (ICD-10) codes. Mild asthma was defined by SNOMED 370218001 or 426979002 and ICD-10 J45.20 or J45.30, whereas moderate-to-severe asthma was defined by SNOMED 427295004 or 426656000 and ICD-10 J45.40 or J45.50. Severity categories were mutually exclusive. Non-asthma controls were participants without EHR or survey-reported asthma, impaired lung function, respiratory or allergic comorbidities, or asthma medication use.

### Genetic Ancestry

Genetic ancestry was assigned by the AoURP using genomic data and Rapid ancestrY Estimation (RYE), a principal component analysis (PCA)-based method trained on reference populations from the Genome Aggregation Database (gnomAD), the Human Genome Diversity Project, and the 1000 Genomes Project. Participants were assigned to the ancestry group with the highest predicted ancestry fraction: European, African, Latino/Admixed, East Asian, South Asian, and Middle Eastern/North African.^12,13^

### Genotyping and Quality Control

DNA extracted from EDTA whole blood was genotyped at the Broad Institute, Johns Hopkins University and the University of Washington using the Illumina Global Diversity Array and harmonized protocols to minimize batch effects. Quality control included assessment of sample swaps, contamination and processing errors, with failing samples excluded. Mean genotype call rate exceeded 98% with no evidence of contamination.^13^ We additionally excluded participants or SNPs with minor allele frequency <1%, Hardy–Weinberg equilibrium *P* < 1 × 10^-6^, outlier heterozygosity > 3 standard deviations above mean, sex discordance, or identity-by-descent ≥0.1.

### Statistical Analysis

Participant characteristics were compared among mild asthma cases, moderate-to-severe asthma cases, and non-asthma controls using chi-square tests for categorical variables and independent *t*-tests for continuous variables. A two-sided *P* < 0.05 was statistically significant.

GWAS were performed using logistic regression under an additive genetic model implemented in PLINK v1.9 for 14,372 mild asthma cases and 7,096 moderate-to-severe asthma cases versus 28,816 controls in pooled multi-ancestry analyses, including participants of European, African, Latino/admixed, East Asian, Middle Eastern/North African, and South Asian ancestries. Ancestry-specific GWAS were subsequently conducted in European (18,356 controls, 8,882 mild, and 4,038 moderate-to-severe cases), African (3,746 controls, 3,032 mild, and 1,859 moderate-to-severe cases), and Latino/admixed (4,985 controls, 2,098 mild, and 1,064 moderate-to-severe cases) participants. Ancestry-specific GWAS results were combined using inverse-variance weighted fixed-effect meta-analysis. Between-ancestry heterogeneity was assessed using Cochran’s *Q* statistic and the *I*² metric. All models were adjusted for age, sex, family income, area deprivation index (ADI), and the first 10 principal components in pooled analyses or the first 10 ancestry-specific principal components in ancestry-specific analyses to account for population stratification.^14,15^ Participants with missing phenotype or covariate data were excluded from the corresponding analyses. Associations are reported as odds ratios (ORs) with 95% confidence intervals (CIs). Genomic coordinates were based on the GRCh38 reference genome. Genome-wide significance was defined as *P* < 5 × 10^-8^. Quantile-quantile plots and genomic inflation factors (λ) were used to assess residual population stratification.

### Expression Quantitative Trait Loci (eQTL) and Pathway Enrichment

Genome-wide significant loci were evaluated for regulatory effects on gene expression in lung tissue using the FIVEx expression quantitative trait loci (eQTL) browser.^16,17^ A 100-kb window centered on each significant SNP was used to identify proximal regulatory variants while minimizing multiple testing and long-range genomic confounding. Candidate genes supported by lung eQTL evidence were subsequently analyzed using gene set over-representation in ConsensusPathDB based on Gene Ontology to identify the enriched biological processes.^18^

## RESULTS

### Characteristics of Study participants

Of the 633,248 AoURP version 8 participants, 340,466 had both EHR and genomic data available. We excluded participants with asthma of undetermined severity, respiratory comorbidities or alternative diagnoses, and missing covariate data. Non-asthma controls were additionally selected using stringent exclusion criteria to minimize phenotypic misclassification (detailed description of the control selection criteria in the Supplementary Methods). Fifteen percent of the eligible participants were randomly held out as an independent dataset for future polygenic risk score (PRS) development and validation. The remaining 85% comprised the final GWAS analytic cohort, including 28,816 non-asthma controls, 14,372 mild asthma cases, and 7,096 moderate-to-severe asthma cases (**Figure 1**).

**Figure 1.**
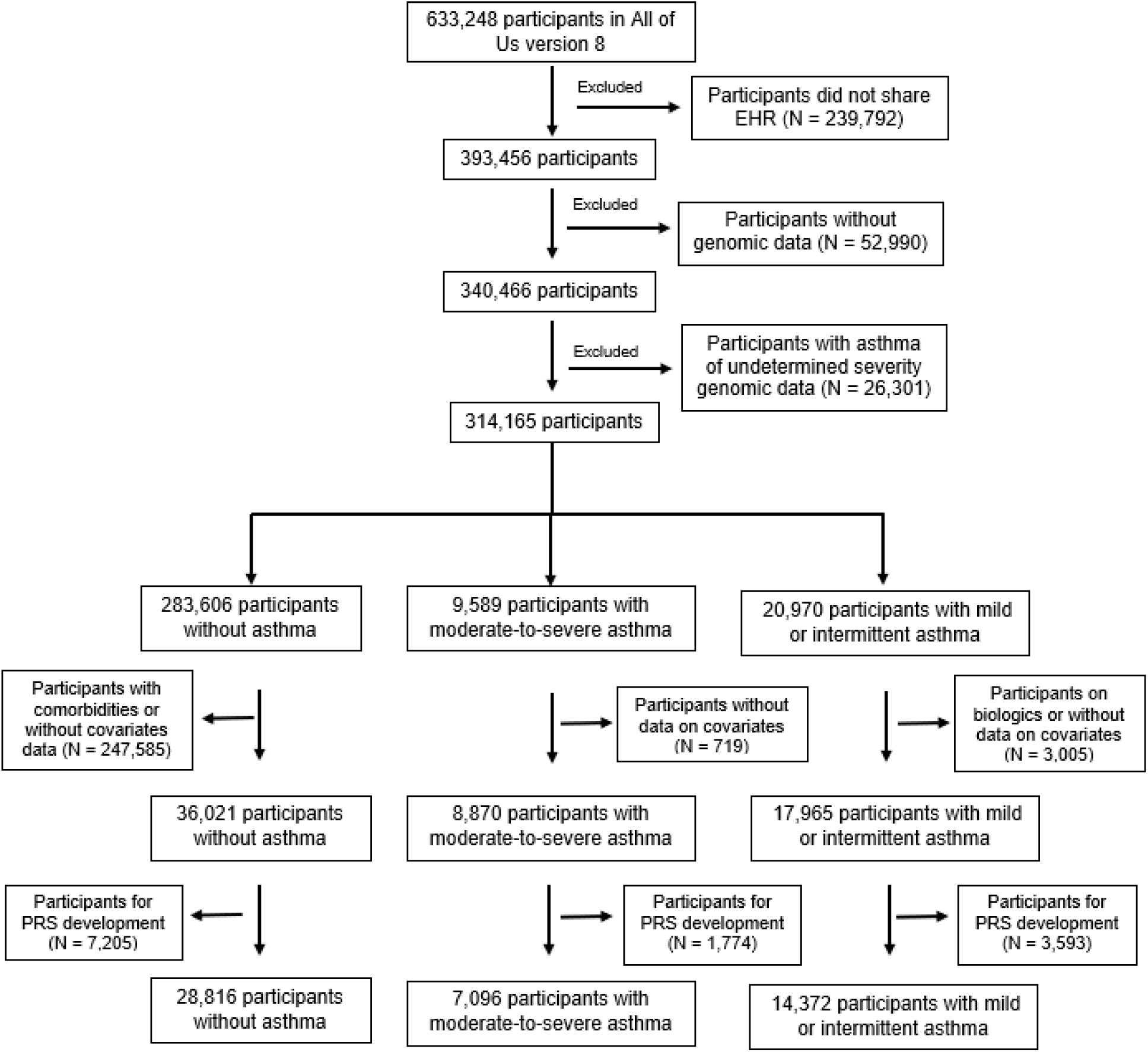
Flowchart of Study Participants included in GWAS Analysis Abbreviations: EHR: electronic health record; PRS: polygenic risk score.

Among the 50,284 participants included in the GWAS, the mean age was 51.9 years, 65.4% were female, the mean annual family income was $65,700, and the mean area deprivation index (ADI) was 31.9. Participants were predominantly of European ancestry (62.2%), followed by African (17.2%) and Latino/admixed (16.2%) ancestries. Compared with non-asthma controls, participants with mild or moderate-to-severe asthma were older, more frequently female or of African ancestry, and had lower family income. Compared participants with mild asthma, those with moderate-to-severe asthma were older, more frequently of African ancestry, had lower family income and higher ADI (**Table 1**).

**Table 1.** Characteristics of study participants, All of Us Research Program (N = 50,284)

| Characteristics | All<br>(N = 50,284) | Controls<br>(N = 28,816) | Asthma |  |  |  |  |
| --- | --- | --- | --- | --- | --- | --- | --- |
|  |  |  | Mild<br>(N = 14,372) |  | Moderate-to-Severe<br>(N = 7,096) |  | P-value <sup>b</sup> |
|  |  |  | Estimate | P-value <sup>a</sup> | Estimate | P-value <sup>a</sup> |  |
| Age at consent, years (mean [SD]) | 51.9 (17.2) | 51.3 (17.7) | 51.9 (16.5) | < 0.001 | 54.1 (15.3) | < 0.001 | < 0.001 |
| Female sex, % | 65.4 | 57.2 | 76.4 | < 0.001 | 76.6 | < 0.001 | 0.74 |
| Genetically inferred ancestry, % |  |  |  | < 0.001 |  | < 0.001 | < 0.001 |
| European | 62.2 | 63.7 | 61.8 |  | 56.9 |  |  |
| African | 17.2 | 13.0 | 21.1 |  | 26.2 |  |  |
| Latino/Admixed | 16.2 | 17.3 | 14.6 |  | 15.0 |  |  |
| Other <sup>c</sup> | 4.4 | 6.0 | 2.5 |  | 1.9 |  |  |
| Family income, per \$1,000 (mean [SD]) | 65.7 (16.5) | 66.0 (16.3) | 65.4 (17.0) | < 0.001 | 64.8 (16.5) | < 0.001 | 0.01 |
| Area Deprivation Index, % (mean [SD]) | 31.9 (6.3) | 32.1 (6.1) | 31.5 (6.5) | < 0.001 | 32.0 (6.5) | 0.17 | < 0.001 |

Abbreviations: SD: standard deviation.
<sup>a</sup> P-values for mild or moderate-to-severe asthma compared to non-asthma controls
<sup>b</sup> P-values for mild compared to moderate-to-severe asthma
<sup>c</sup> "Other" include East Asian, South Asian, and Middle Eastern/North African ancestries
P-values computed using independent t-tests for continuous variables (age at consent, family income and area deprivation index) or chi-square for the categorical variables (sex and genetically inferred ancestry)

### GWAS of Mild Asthma versus No Asthma

The pooled multi-ancestry GWAS identified 93 genome-wide significant variants (λ: 1.19), while the fixed-effect meta-analysis identified 104 genome-wide significant variants (λ: 1.12) (**Figure 2**; **Supplementary Tables S1 and S2**). In both analyses, the strongest associations localized to the chromosome 2q12 asthma susceptibility locus encompassing *IL1RL1, IL18R1, IL18RAP*, and *SLC9A4*, followed by the HLA class II region on chromosome 6 involving *HLA-DQA1, HLA-DQB1*, and *HLA-DRB1*. Additional genome-wide significant associations were observed near *IL33* (*GTF3AP1-IL33*), *LINC02757-EMSY*, *SMAD3*, *IKZF3*, and *GSDMB*. Lung eQTL implicated *IL1RL1, IL18R1, IL18RAP, SLC9A4, IL33, LRRC32, SMAD3, IKZF3, GSDMB, GRB7*, and *MIEN1* as candidate target genes (**Figures 2 and 3; Supplementary Tables S1 and S2**).

**Figure 2:**
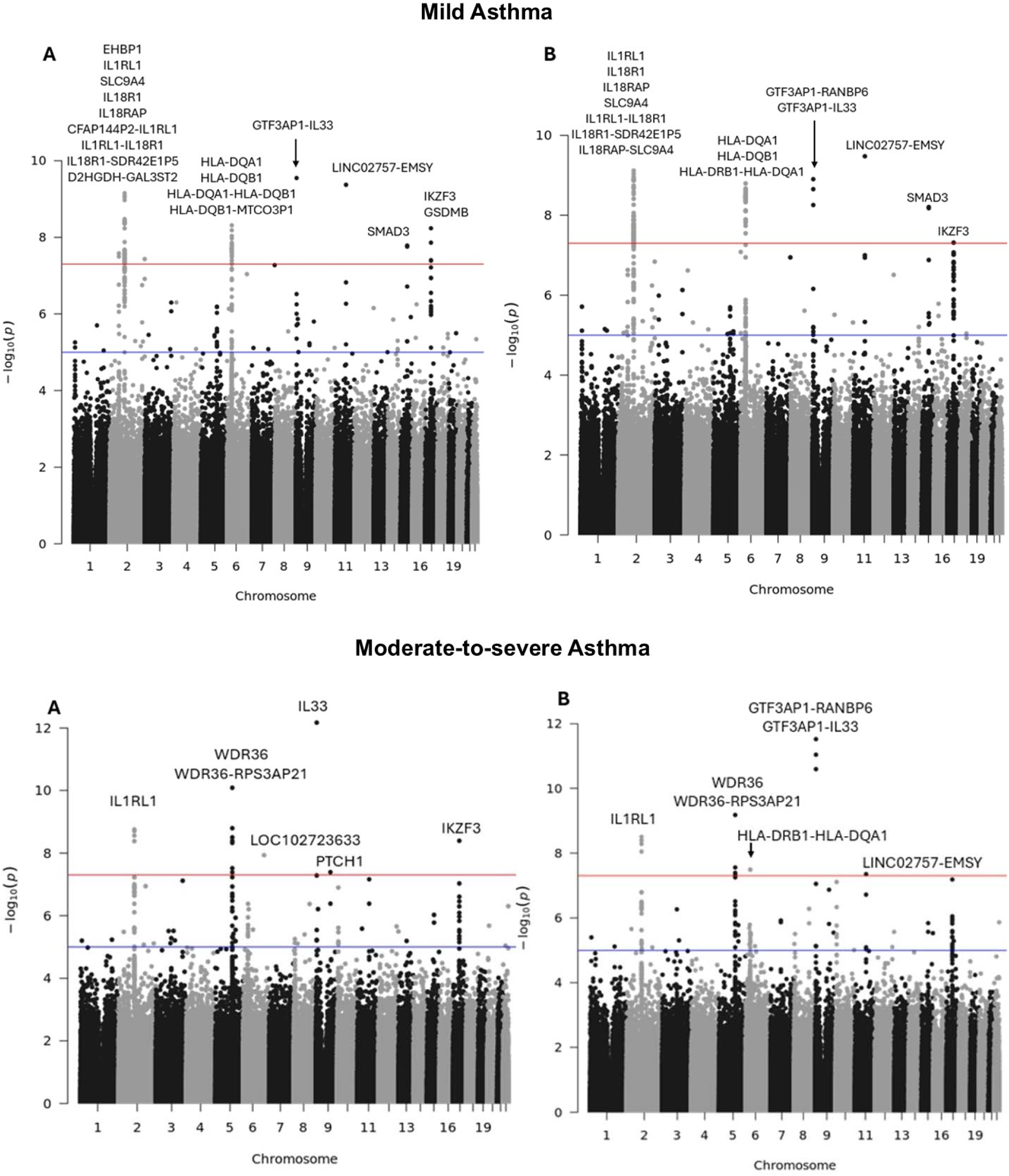
Manhattan Plots for Pooled and Meta-Analyses Multi-Ancestry GWAS of Mild and Moderate-to-severe Asthma Manhattan plots from pooled (A) and fixed-effect meta-analyses (B) of mild asthma (top) and moderate-to-severe asthma (bottom). The red horizontal line indicates the genome-wide significance threshold (*P* < 5 × 10^-8^). The blue horizontal line indicates the suggestive significance threshold (*P* < 1 × 10^-5^).

**Figure 3:**
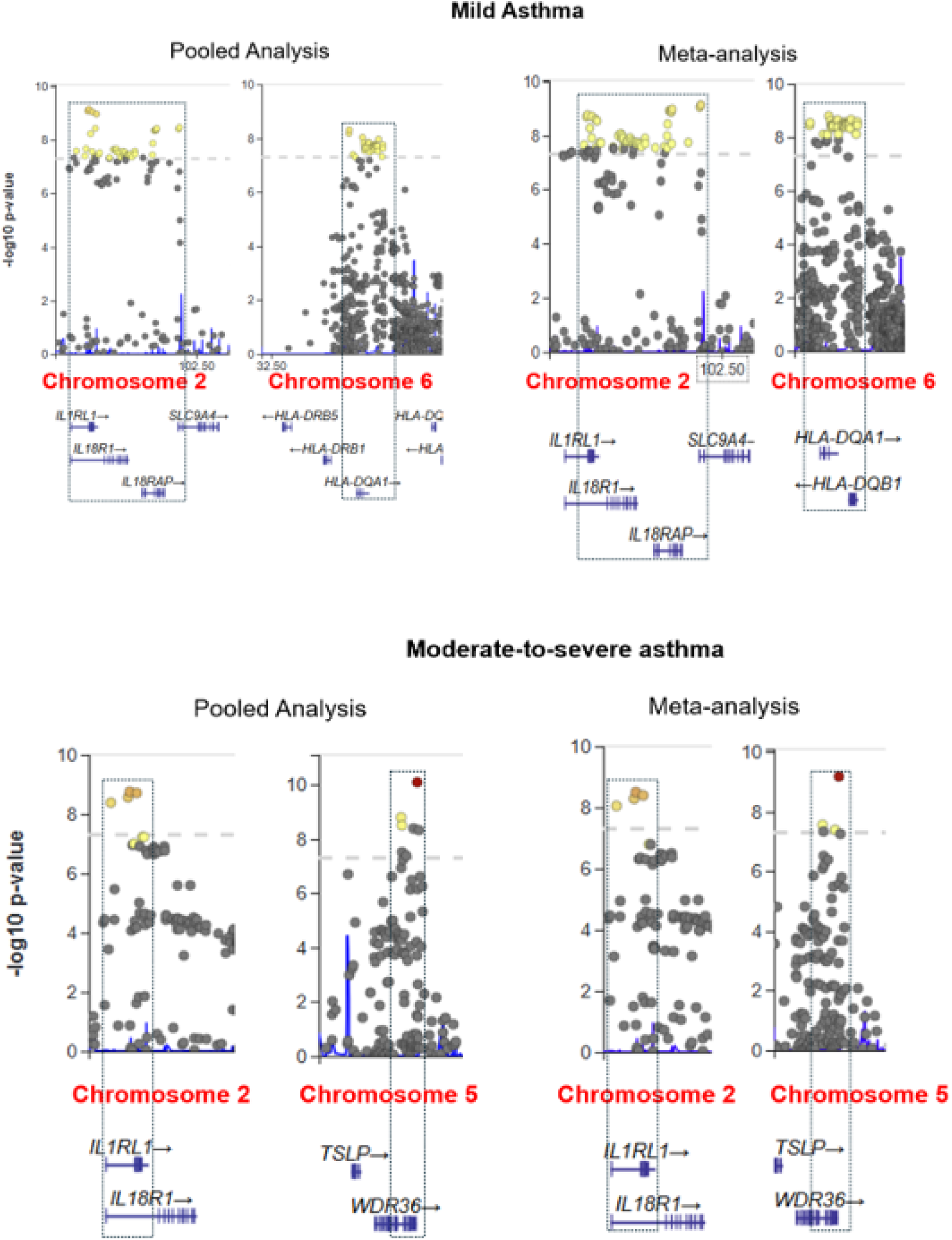
Regional association plots of the major susceptibility loci in mild and moderate-to-severe asthma The figure shows loci on chromosome 2 (*IL1RL1* and *IL18R1*) being shared across mild and moderate-to-severe asthma, with mild asthma additionally including loci on *IL18RAP* and *SLC9A4*. Mild asthma-spe ific loci included *HLA-DQA1* and *HLA-DQB1* on chromosome 6, whereas moderate-to-severe asthma-specific locus included *WDR36* on chromosome 5. Points colored according to linkage disequilibrium (LD) r^2^ with lead SNP based on the selected LocusZoom reference populations. **Red dots** = LD r² with lead SNP ≥ 0.8. **Orange dots**= LD r² with lead SNP ≥ 0.6 and < 0.8. **Yellow dots** = LD r² with lead SNP ≥ 0.4 and < 0.6.

The Manhattan plots from the ancestry-specific GWAS of mild asthma are presented in **Supplementary Figures S1-S3.**

### GWAS of Moderate-to-Severe Asthma versus No Asthma

The pooled multi-ancestry GWAS and fixed-effect meta-analysis identified 15 (λ: 1.19) and 14 (λ: 1.11) genome-wide significant variants, respectively (**Figure 2; Supplementary Tables S3 and S4**). In both analyses, the strongest associations mapped to *IL33* locus and the *WDR36* locus, with lung eQTL implicating *WDR36*, *TSLP*, and *CAMK4*. We also found genome-wide significant associations at *IL1RL1*, where lung eQTL implicated *IL1RL1*, *IL18R1*, and *IL18RAP*.

Additional significant variants differed between analyses. The pooled GWAS identified loci near *LOC102723633*, *PTCH1*, and *IKZF3*. Lung eQTL further implicated *GSDMB*, *IKZF3*, *GRB7*, *FANCC*, *PTCH1*, and *MIEN1*. In contrast, the fixed-effect meta-analysis identified significant variants within the HLA class II region and the *LINC02757*-*EMSY* region and lung eQTL implicated *HLA-DRB1*, *HLA-DQA1*, *HLA-DRB5*, *HLA-DQB1*, *HLA-DQB1-AS1*, and *LRRC32*. We observed dense clusters of significant variants at the *IL1RL1* and *WDR36* loci in both analyses (**Figure 3**).

The Manhattan plots from the ancestry-specific GWAS of moderate-to-severe asthma are presented in **Supplementary Figures S1-S3**.

### Comparison of Genetic Architectures in Mild and Moderate-to-Severe Asthma

#### Genome-wide Association Analyses

Shared genome-wide significant loci between mild and moderate-to-severe asthma included *IL1RL1*, *IKZF3*, *HLA-DRB1-HLA-DQA1*, *GTF3AP1-IL33*, and *LINC02757-EMSY* regions (Table 2). The *IL33* locus showed a stronger association with moderate-to-severe asthma than mild asthma, with higher odds ratios (1.16-1.17 vs. 1.11-1.12) and more significant *P* values (6.69 × 10^-13^ to 2.55 x 10^-11^ vs. 2.83 x 10^-10^ to 1.25 x 10^-9^), suggesting a severity gradient in genetic risk (**Supplementary Tables S1-S4**).

**Table 2:** Shared and specific genome-wide significant loci from pooled and meta-analyses across mild and moderate-to-severe asthma.

| Chr | Locus | Mild Asthma |  | Moderate-to-Severe Asthma |  | Shared or Specific |
| --- | --- | --- | --- | --- | --- | --- |
|  |  | Pooled | Meta-analysis | Pooled | Meta-analysis |  |
| 2 | *EHBP1* | ■ |  |  |  | Mild |
| 2 | *CFAP144P2-IL1RL1* | ■ | ■ |  |  | Mild |
| 2 | *IL1RL1* | ■ | ■ | ■ | ■ | Shared |
| 2 | *IL1RL1-IL18R1* | ■ | ■ |  |  | Mild |
| 2 | *IL18R1* | ■ | ■ |  |  | Mild |
| 2 | *IL18R1-SDR42E1* | ■ | ■ |  |  | Mild |
| 2 | *IL18RAP* | ■ | ■ |  |  | Mild |
| 2 | *IL18RAP-SLC9A4* |  | ■ |  |  | Mild |
| 2 | *D2HGDH-GAL3ST2* | ■ |  |  |  | Mild |
| 5 | *WDR36* |  |  | ■ | ■ | Moderate-to-severe |
| 5 | *WDR36-RPS3AP21* |  |  | ■ | ■ | Moderate-to-severe |
| 6 | *LOC102723633* |  |  | ■ |  | Moderate-to-severe |
| 6 | *HLA-DRB1-HLA-DQA1* |  | ■ |  | ■ | Shared |
| 6 | *HLA-DQA1* | ■ |  |  |  | Mild |
| 6 | *HLA-DQA1-HLA-DQB1* | ■ |  |  |  | Mild |
| 6 | *HLA-DQB1* | ■ |  |  |  | Mild |
| 6 | *HLA-DQB1-MTCO3P1* | ■ |  |  |  | Mild |
| 9 | *PTCH1* |  |  | ■ |  | Moderate-to-severe |
| 9 | *GTF3AP1-IL33* | ■ |  | ■ | ■ | Shared |
| 9 | *GTF3AP1-RANBP6* |  | ■ | ■ |  | Shared |
| 11 | *LINC02757-EMSY* | ■ |  | ■ | ■ | Shared |
| 15 | *SMAD3* | ■ |  |  |  | Mild |
| 17 | *IKZF3* | ■ |  | ■ |  | Shared |
| 17 | *GSDMB* | ■ |  |  |  | Mild |

Chr = chromosome; ■ = Locus present in GWAS analysis. Blue shade indicates mild-specific loci. Red shade indicates moderate-to-severe-specific loci.

Mild asthma demonstrated broader association signals across both the chromosome 2q12 and HLA class II regions. At chromosome 2q12, the associations extended beyond the shared *IL1RL1* locus to include *IL18R1, IL18RAP*, and *SLC9A4*, together with the *IL1RL1-IL18R1, IL18RAP-SLC9A4*, and *IL18R1-SDR42E1P5* intergenic regions. Within the HLA class II region, we observed additional associations at *HLA-DQA1, HLA-DQB1, HLA-DQA1-HLA-DQB1*, and *HLA-DQB1-MTCO3P1*. Mild asthma-specific loci included *EHBP1*, *D2HGDH*-*GAL3ST2*, and *SMAD3*, whereas moderate-to-severe asthma-specific loci included *WDR36*, *WDR36*-*RPS3AP21*, and *PTCH1* (**Table 2**).

#### Lung eQTL Analyses

The shared lung eQTL candidate genes were *IL1RL1, IL18R1, IL18RAP, IL33, LRRC32,* and *MIEN1* and HLA class II genes (*HLA-DRB1, HLA-DQA1, HLA-DQB1* and *HLA-DRB5*). Mild asthma-specific lung eQTL candidate genes included HLA class II genes (*HLA-DQA2, HLA-DQB2* and *HLA-DRA*), the 17q12-21 asthma susceptibility genes (*GSDMB, GSDMA, ORMDL3* and *PSMD3*) and *SMAD3*. In contrast, moderate-to-severe asthma-specific candidate genes included *TSLP, WDR36, CAMK4, PTCH1,* and *FANCC* (**Table 3**).

**Table 3:** Shared and specific lung eQTL genes from pooled and meta-analyses across mild and moderate-to-severe asthma.

| Chr | eQTL Genes | Mild Asthma |  | Moderate-to-Severe Asthma |  | Shared or Specific |
| --- | --- | --- | --- | --- | --- | --- |
|  |  | Pooled | Meta-analysis | Pooled | Meta-analysis |  |
| 2 | *IL1RL1, IL18R1, IL18RAP* | ■ | ■ | ■ | ■ | Shared |
| 2 | *IL1RL2* | ■ | ■ |  |  | Mild |
| 2 | *D2HGDH, DTYMK, PDCD1, NEU4, GAL3ST2* | ■ |  |  |  | Mild |
| 5 | *WDR36, TSLP, CAMK4* |  |  | ■ | ■ | Moderate-to-severe |
| 6 | *HLA-DQA1, HLA-DQB1, HLA-DRB1, HLA-DRB5* | ■ | ■ |  | ■ | Shared |
| 6 | *HLA-DQB1-AS1* |  | ■ |  | ■ | Shared |
| 6 | *HLA-DQA2, HLA-DQB2, HLA-DRA* | ■ |  |  |  | Mild |
| 9 | *IL33* | ■ | ■ | ■ | ■ | Shared |
| 9 | *PTCH1, FANCC* |  |  | ■ |  | Moderate-to-severe |
| 11 | *LRRC32* | ■ | ■ |  | ■ | Shared |
| 15 | *SMAD3, IQCH, AAGAB* | ■ | ■ |  |  | Mild |
| 17 | *IKZF3, GRB7, MIEN1* | ■ | ■ | ■ |  | Shared |
| 17 | *GSDMB* | ■ |  | ■ |  | Shared |
| 17 | *GSDMA, ORMDL3, LRRC3C, PSMD3* | ■ |  |  |  | Mild |

Chr = chromosome; ■ = gene present in lung eQTL for SNPs significant in analysis. Blue shade indicates mild-specific lung eQTL gene. Red shade indicates moderate-to-severe-specific lung eQTL gene.

#### Pathway Enrichment Analyses

Pathway analyses identified shared enrichment of immune responses and cytokine signaling across asthma severities. Mild asthma was characterized by enrichment of TGF-β signaling, T-cell differentiation, and IL-1-mediated signaling pathways. In contrast, moderate-to-severe asthma showed stronger enrichment of IL-6, IL-10, and IL-13 production, myeloid cell differentiation, and cellular responses to reactive oxygen species (**Table 4**).

**Table 4:** Summary results of gene set over representation analysis on lung eQTL genes from significant SNPs in pooled and meta-analyses.

| Biological processes | Mild asthma |  | Moderate-to-severe asthma |  |
| --- | --- | --- | --- | --- |
|  | Pooled | Meta-analysis | Pooled | Meta-analysis |
| Regulation of interleukin-5 production | ■ | ■ | ■ | ■ |
| Interleukin-33-mediated signaling pathway | ■ | ■ | ■ | ■ |
| Regulation of interleukin-4 production | ■ | ■ |  | ■ |
| Antigen processing and presentation via MHC class II | ■ | ■ |  | ■ |
| Cytokine signaling pathways | ■ | ■ | ■ | ■ |
| Interleukin-18-mediated signaling pathway | ■ | ■ | ■ | ■ |
| T cell mediated immunity | ■ | ■ |  | ■ |
| B cell mediated immunity |  | ■ |  | ■ |
| Macrophage activation | ■ | ■ | ■ | ■ |
| NF-kappa B activation |  | ■ | ■ | ■ |
| Chemokine production | ■ | ■ | ■ | ■ |
| Immune regulatory pathways | ■ | ■ | ■ | ■ |
| Interferon-gamma-mediated signaling pathway | ■ | ■ | ■ | ■ |
| Transforming growth factor beta signaling | ■ | ■ |  |  |
| Leukocyte adhesion and cell-cell interactions | ■ |  |  |  |
| T cell differentiation and function | ■ |  |  |  |
| Leukocyte differentiation and hematopoiesis | ■ |  |  |  |
| Interleukin-1-mediated signaling pathway | ■ |  |  |  |
| Leukocyte apoptotic process | ■ |  |  |  |
| Ceramide metabolic process | ■ |  |  |  |
| Protein modification by small protein removal | ■ |  |  |  |
| Negative regulation of defense response |  | ■ |  |  |
| Regulation of interleukin-10 production |  |  |  | ■ |
| Regulation of interleukin-6 production |  |  | ■ | ■ |
| Regulation of interleukin-13 production |  |  | ■ | ■ |
| Myeloid leukocyte differentiation |  |  |  | ■ |
| Cellular response to reactive oxygen species |  |  | ■ |  |
| Regulation of macromolecule biosynthetic process |  |  | ■ |  |

Blue shade indicates mild-specific biological process. Red shade indicates moderate-to-severe-specific biological process

## DISCUSSION

Our GWAS of 50,284 participants represents the first multi-ancestry study to directly compare the genetic architectures of mild and moderate-to-severe asthma. We identified a shared genetic core and distinct severity-specific loci. Lung eQTL implicated additional shared candidate genes (*IL18R1, IL18RAP,* and *GSDMB*). In pathway analysis, TGF-β signaling and T-cell differentiation were enriched in mild asthma. Moderate-to-severe asthma was characterized by IL-6, IL-10, and IL-13 production, myeloid cell differentiation, and cellular responses to reactive oxygen species.

### Shared Genetic Architecture

Five loci reached genome-wide significance in both mild and moderate-to-severe asthma: *IL1RL1*, *IKZF3* and the *HLA-DRB1-HLA-DQA1*, *GTF3AP1-IL33*, and *LINC02757-EMSY* intergenic regions. *IL1RL1* encodes ST2, the receptor for the epithelial alarmin IL-33, which promotes type 2 inflammation and eosinophilic airway disease through induction of IL-4, IL-5, and IL-13.^19^ Lung eQTL at this locus implicated *IL18R1* and *IL18RAP*, supporting a shared role for IL-33 and IL-18 signaling across asthma severities. Although the *GTF3AP1-IL33* locus was shared, it showed stronger associations in moderate-to-severe asthma, consistent with higher pulmonary IL-33 expression in severe disease.^20^

Lung eQTL further implicated *LRRC32*, *IKZF3*, and HLA class II genes (*HLA-DRB1, HLA-DQA1, HLA-DQB1,* and *HLA-DRB5*). *LRRC32*, a regulator of TGF-β activation, has been linked to immune homeostasis and airway remodeling.^21^ *IKZF3* regulates lymphocyte development, IgE class switching, and Th2 differentiation and has been implicated in eosinophilic airway inflammation and asthma susceptibility.^22^ Together with HLA-mediated antigen presentation, these findings support a shared genetic framework centered on adaptive immune regulation.

Consistent with these findings, pathway analyses identified shared enrichment of IL-33 and IL-18 signaling, antigen presentation, T-cell immunity, interferon-γ signaling, NF-κB activation, and inflammatory response pathways. Overall, this suggests that shared genetic architecture reflects upstream immune mechanisms involving epithelial alarmin signaling, antigen presentation, and adaptive immune regulation, while severity-specific loci may influence downstream pathways underlying disease severity.

### Mild Asthma-Specific Genetic Architecture

Mild asthma exhibited a broader genetic architecture than moderate-to-severe asthma, with associations at *EHBP1, D2HGDH-GAL3ST2, SMAD3, GSDMB*, and multiple HLA class II genes. Although adaptive immune pathways, including antigen presentation and T-cell activation, were shared across asthma severities, mild asthma showed additional enrichment of T-cell differentiation, TGF-β regulation, and lymphocyte homeostasis, consistent with a greater contribution of allergen sensitization and immune regulation.

The broader HLA class II signal in mild asthma supports a role for antigen presentation in atopic sensitization and antigen-specific IgE responses.^23^ The consistent protective associations across HLA loci with odds ratios ranging from 0.87 to 0.92 further suggest that specific HLA class II variants may be associated with reduced susceptibility to asthma through altered antigen presentation and adaptive immune activation.^23^ The *SMAD3* locus was identified as a mild asthma-specific lung eQTL together with *IQCH* and *AAGAB*. *SMAD3* is a central mediator of TGF-β signaling that regulates immune tolerance and regulatory T-cell differentiation.^24^ Combined with the shared *LRRC32* eQTL and enrichment of TGF-β regulatory pathways, these findings suggest that TGF-β signaling in mild asthma may reflect immune regulation rather than the pro-fibrotic functions associated with severe disease. Within the 17q12-21 asthma locus, lung eQTL implicated *GSDMA, ORMDL3,* and *PSMD3* specifically in mild asthma, whereas *GSDMB* was shared across the phenotypes. This region is strongly associated with childhood-onset and atopic asthma and further supports a predominance of adaptive immune mechanisms in mild asthma.^25^

Consistent with the GWAS and lung eQTL findings, pathway analyses demonstrated enrichment of T-helper cell differentiation, memory T-cell differentiation, leukocyte differentiation, IL-1 signaling, and TGF-β regulation. Overall, these findings suggest that mild asthma is characterized by enhanced antigen presentation, adaptive immune activation, and TGF-β mediated immunoregulation.

### Moderate-to-Severe Asthma-Specific Genetic Architecture

Moderate-to-severe asthma was characterized by associations at *WDR36, WDR36-RPS3AP21,* and *PTCH1*, along with enrichment of innate inflammatory and oxidative stress pathways, supporting mechanisms previously implicated in steroid-resistant severe asthma.

Variants at *WDR36* and *WDR36-RPS3AP21* acted as lung eQTL for *TSLP, WDR36,* and *CAMK4*. *TSLP* is an epithelial alarmin that initiates type 2 immune responses and is a key therapeutic target in severe asthma.^26^ Lung eQTL also implicated *CAMK4*, which suppresses regulatory T-cell function through reduced *FOXP3* expression, suggesting that impaired immune regulation may contribute to persistent airway inflammation.^27^ The *PTCH1* locus has previously been associated with impaired lung function.^28^ As the primary receptor of the Hedgehog signaling pathway, *PTCH1* regulates epithelial repair and airway remodeling, and Hedgehog inhibition attenuates airway inflammation in experimental asthma.^29^ Lung eQTL additionally implicated *FANCC* involved in cellular stress and DNA damage responses.^29^

Consistent with these GWAS and lung eQTL findings, pathway analyses identified enrichment of IL-13, IL-6, and IL-10 production, myeloid cell differentiation, and cellular responses to reactive oxygen species. IL-13 enrichment supports enhanced type 2 inflammation, whereas IL-6 is consistent with innate immune activation and steroid-resistant inflammation.^30,31^ Enrichment of IL-10 pathways may reflect compensatory anti-inflammatory responses that are insufficient to control persistent airway inflammation.^32^ Together, these findings suggest that moderate-to-severe asthma is characterized by epithelial alarmin signaling, innate inflammatory amplification, oxidative stress, and airway remodeling, distinguishing it from the predominantly adaptive immune profile of mild asthma.

### Implications

Our findings have potential therapeutic implications, particularly for moderate-to-severe asthma. First, the identification of *TSLP* as a lung eQTL supports the importance of epithelial alarmin signaling and provides genetic support for the clinical efficacy of anti-*TSLP* therapy.^33^ Second, enrichment of IL-13 production pathways reinforces the central role of type 2 inflammation. However, the limited efficacy of anti-IL-13 therapies suggests that moderate-to-severe asthma includes multiple inflammatory endotypes involving IL-4, IL-5, innate immune activation, and epithelial alarmin signaling.^34^ Third, enrichment of IL-6 production pathways implicates innate-adaptive immune crosstalk and steroid-resistant inflammation, highlighting a potential therapeutic gap because no IL-6-targeted therapies are currently approved for asthma.^31^ Nevertheless, the anti-IL-6 tocilizumab has been explored off-label in severe asthma and the PrecISE trial is evaluating clazakizumab in a biomarker-enriched population.^35^ Finally, if replicated, the *PTCH1/FANCC* locus implicating Hedgehog signaling may represent a novel therapeutic target for moderate-to-severe asthma.

### Limitations and Strengths

Several limitations should be considered. Mild and moderate-to-severe asthma were defined using ICD-10 and SNOMED codes, which may have resulted in phenotypic misclassification because disease severity can change over time and treatment intensity is not fully captured by electronic health records. Although our multi-ancestry design increased generalizability, ancestry-specific analyses had limited statistical power to detect population-specific associations, particularly in non-European populations.^36–38^ Our functional interpretation was based on lung eQTL and gene set over-representation analyses rather than experimental validation, and the novel findings, including the *PTCH1/FANCC* locus, require replication in independent cohorts.

Despite these limitations, our study has several strengths. This is the first multi-ancestry GWAS to directly compare the genetic architectures of mild and moderate-to-severe asthma. The large, ancestrally diverse cohort extends previous studies conducted predominantly in European populations and allowed for both pooled and ancestry-specific analyses. Integration of GWAS, lung eQTL, and pathway enrichment analyses provided complementary functional evidence supporting the biological interpretation of associated loci. Moreover, the concordance between pooled and fixed-effect meta-analysis findings strengthens the robustness of our conclusions.

## Conclusions

In the present multi-ancestry analyses, mild and moderate-to-severe asthma shared a common genetic core centered on the *IL33-IL1RL1* axis but diverged in severity-specific biological mechanisms. Mild asthma was characterized by adaptive immune regulation, whereas moderate-to-severe asthma exhibited epithelial alarmin signaling and innate inflammatory pathways. These findings provide a mechanistic framework for asthma heterogeneity and precision medicine. Future ancestry-specific analyses are warranted to determine whether the severity-specific genetic and molecular mechanisms are consistent or vary across ancestry.

## Data Availability

This present study used publicly available data available through the workbench of the NIH All of Us Research Program.

https://www.nih.gov/allofus

