## Supplementary Material for "Multi-Ancestry Genome-wide Association Analyses Identify Shared and Specific Genetic Architecture in Mild and Moderate-to-Severe Asthma"

Angelico Mendy,^1^ Yadu Gautam,^2^ Bradley H. Rosen,^2^ Joseph Castlen,^3^ Michael B. Fessler,^4^ Darryl C. Zeldin,^4^ Peter S. Thorne,^5^ Tesfaye B. Mersha^2^

*^1^Department of Epidemiology and Population Health, School of Public Health, Louisiana State University Health Sciences Center, New Orleans, Louisiana, USA*

*^2^ Division of Pulmonary, Critical Care, Sleep, and Occupational Medicine, Department of Medicine, Indiana University School of Medicine, Indianapolis, Indiana, USA*

*^3^ Division of Pulmonary, Allergy, and Sleep, Department of Pediatrics, Indiana University School of Medicine and Riley Hospital for Children, Indianapolis, Indiana, USA*

*^4^ Division of Intramural Research, National Institute for Environmental Health Sciences, National Institutes of Health, Research Triangle Park, North Carolina, USA*

*^5^ Department of Occupational and Environmental Health, University of Iowa, Iowa City, Iowa, USA*

**Supplementary Methods**

To minimize phenotypic misclassification, participants were excluded from the non-asthma control group if they had evidence suggestive of asthma or related respiratory disease based on diagnoses, pulmonary function, medication use, clinical observations, or self-report. The specific exclusion criteria included: 1) diagnoses of asthma, wheezing, allergic respiratory diseases (e.g., allergic rhinitis, allergic cough, allergic conjunctivitis), atopic disorders (e.g., atopic dermatitis, IgE-mediated allergic disease, urticaria), chronic sinusitis, or other allergic conditions. 2) evidence of airflow obstruction on pulmonary function testing, defined as FEV₁ ≤ 80% predicted, FEV₁/FVC ≤ 80%, FVC ≤ 80% predicted, or FEF_25-75_ ≤65% predicted. 3) use of medications for obstructive airway disease or asthma, including inhaled corticosteroids, short- and long-acting β₂-agonists, muscarinic antagonists, leukotriene receptor antagonists, methylxanthines, anti-IgE therapy, anti-IL-5/IL-5 receptor biologics, anti-IL-4Rα therapy, oxygen therapy, or long-term systemic immunosuppressive or corticosteroid therapy. 4) asthma-related clinical observations, including documented asthma control assessments, hospitalization for bronchitis or asthma, long-term inhaled or systemic corticosteroid use, or a documented personal history of respiratory disease. 5) self-reported asthma or allergies in survey responses.

**Supplementary Table S1**: Genome-wide significant loci and lung eQTL candidate genes identified in the pooled GWAS of mild asthma

| **Position** | **SNPs** |  | **OR** | **P** | **Gene** | **Lung eQTL** |
| --- | --- | --- | --- | --- | --- | --- |
| chr2:62904596 | rs721048 | Intron | 1.13 | 2.62 _x_ 10^-08^ | *EHBP1* | - |
| chr2:62907744 | rs2710646 | Intron | 1.12 | 3.17 _x_ 10^-08^ |  | - |
| chr2:102265892 | rs78545931 | Intergenic | 0.89 | 4.34 _x_ 10^-08^ | *CFAP144P2-IL1RL1* | *IL1RL1, IL1RL2, IL18R1* |
| chr2:102275879 | rs60227565 | Intergenic | 0.87 | 2.39 _x_ 10^-08^ |  |  |
| chr2:102316102 | rs950880 | Intron | 1.09 | 4.60 _x_ 10^-08^ | *IL1RL1* | *IL1RL1, IL18R1, IL18RAP* |
| chr2:102319699 | rs72823641 | Intron | 0.87 | 2.58 _x_ 10^-08^ |  |  |
| chr2:102332701 | rs12479210 | Intron | 1.09 | 3.92 _x_ 10^-08^ |  |  |
| chr2:102334362 | rs13019081 | Intron | 1.09 | 2.10 _x_ 10^-08^ |  |  |
| chr2:102337157 | rs3771180 | Intron | 0.88 | 8.24 _x_ 10^-10^ |  |  |
| chr2:102338622 | rs13408661 | Intron | 0.88 | 7.06 _x_ 10^-10^ |  |  |
| chr2:102341256 | rs1420101 | Intron | 1.10 | 5.82 _x_ 10^-09^ |  |  |
| chr2:102343547 | rs12905 | Intron | 1.11 | 2.73 _x_ 10^-08^ |  |  |
| chr2:102343750 | rs3771175 | Intron | 0.88 | 8.92 _x_ 10^-10^ |  |  |
| chr2:102346612 | rs6543119 | Intron | 1.09 | 4.39 _x_ 10^-08^ |  |  |
| chr2:102348282 | rs13017455 | Intron | 1.09 | 4.02 _x_ 10^-08^ |  |  |
| chr2:102348401 | rs55927292 | Intron | 1.11 | 3.67 _x_ 10^-09^ |  |  |
| chr2:102348872 | rs17027006 | Intron | 1.10 | 4.59 _x_ 10^-08^ |  |  |
| chr2:102350089 | rs10197862 | Intron | 0.88 | 1.07 _x_ 10^-09^ |  |  |
| chr2:102353347 | rs67723747 | Intergenic | 1.10 | 3.46 _x_ 10^-08^ | *IL1RL1-IL18R1* |  |
| chr2:102354705 | rs56386507 | Upstream | 1.10 | 2.99 _x_ 10^-08^ | *IL18R1* |  |
| chr2:102369352 | rs3771172 | Intron | 1.10 | 4.78 _x_ 10^-08^ |  |  |
| chr2:102369490 | rs3771171 | Intron | 1.10 | 4.88 _x_ 10^-08^ |  |  |
| chr2:102369694 | rs2160202 | Intron | 1.10 | 4.58 _x_ 10^-08^ |  |  |
| chr2:102375619 | rs2270298 | Intron | 1.10 | 2.25 _x_ 10^-08^ |  |  |
| chr2:102378424 | rs17027037 | Intron | 1.10 | 2.25 _x_ 10^-08^ |  |  |
| chr2:102380345 | rs11683700 | Intron | 1.10 | 2.62 _x_ 10^-08^ |  |  |
| chr2:102380412 | rs3821203 | Intron | 1.10 | 2.16 _x_ 10^-08^ |  |  |
| chr2:102381273 | rs11465633 | Intron | 1.10 | 2.91 _x_ 10^-08^ |  |  |
| chr2:102384942 | rs1035130 | Intron | 1.10 | 3.01 _x_ 10^-08^ |  |  |
| chr2:102386805 | rs2241116 | Intron | 1.10 | 4.36 _x_ 10^-08^ |  |  |
| chr2:102389927 | rs4851570 | Intron | 1.10 | 2.65 _x_ 10^-08^ |  |  |
| chr2:102391232 | rs1680552504 | Intron | 1.10 | 3.59 _x_ 10^-08^ |  |  |
| chr2:102392509 | rs66919607 | Intron | 1.10 | 4.22 _x_ 10^-08^ |  |  |
| chr2:102392952 | rs11679146 | Intron | 0.87 | 2.83 _x_ 10^-08^ |  |  |
| chr2:102394070 | rs2287035 | Intron | 1.10 | 4.78 _x_ 10^-08^ |  |  |
| chr2:102396214 | rs17027071 | Intron | 1.10 | 3.43 _x_ 10^-08^ |  |  |
| chr2:102397842 | rs1135354 | 3’UTR | 1.10 | 3.57 _x_ 10^-08^ |  |  |
| chr2:102399458 | rs17027087 | Intergenic | 1.10 | 3.52 _x_ 10^-08^ | *IL18R1-SDR42E1P5* |  |
| chr2:102401617 | rs3732123 | Intergenic | 1.10 | 3.35 _x_ 10^-08^ |  |  |
| chr2:102403322 | rs55742125 | Intergenic | 1.10 | 4.28 _x_ 10^-08^ |  |  |
| chr2:102407871 | rs55883125 | Intergenic | 1.10 | 2.61 _x_ 10^-08^ |  |  |
| chr2:102408278 | rs4851006 | Intergenic | 1.10 | 3.37 _x_ 10^-08^ |  |  |
| chr2:102420217 | rs3771156 | Intron | 1.10 | 4.59 _X_ 10^-08^ | *IL18RAP* | *IL18RAP* |
| chr2:102434402 |  |  | 1.10 | 3.61 _X_ 10^-08^ |  |  |
| chr2:102435098 | rs4851582 | Intron | 1.10 | 1.33 _X_ 10^-08^ |  |  |
| chr2:102436842 | rs58883541 | Intron | 1.11 | 4.41 _X_ 10^-09^ |  |  |
| chr2:102438960 | rs17027166 | Intron | 1.11 | 4.39 _X_ 10^-09^ |  |  |
| chr2:102439201 | rs55645612 | Intron | 1.11 | 4.18 _X_ 10^-09^ |  |  |
| chr2:102439636 | rs56166614 | Intron | 1.11 | 4.19 _X_ 10^-09^ |  |  |
| chr2:102440699 | rs17027179 | Intron | 1.11 | 3.69 _X_ 10^-09^ |  |  |
| chr2:102473668 | rs17027255 | 5’UTR | 1.11 | 3.90 _X_ 10^-09^ | *SLC9A4* | *IL1RL1, IL18R1, IL18RAP* |
| chr2:102475081 | rs17027258 | Intron | 1.11 | 3.34 _X_ 10^-09^ |  |  |
| chr2:241771867 | rs62192043 | Intergenic | 0.91 | 3.69 _X_ 10^-08^ | *D2HGDH-GAL3ST2* | *D2HGDH, DTYMK, PDCD1, NEU4, GAL3ST2* |
| chr6:32616804 | rs73730372 | Intergenic | 0.88 | 6.54 _X_ 10^-09^ | *HLA-DRB1-HLA-DQA1* | *HLA-DRB1, HLA-DQA1, HLA-DRB5, HLA-DQB1, HLA-DQB2, HLA-DQA2, HLA-DRA* |
| chr6:32617294 | rs113243185 | Intergenic | 0.87 | 4.86 _X_ 10^-09^ |  |  |
| chr6:32622991 | rs34831921 | Intergenic | 0.87 | 3.97 _X_ 10^-08^ |  |  |
| chr6:32626326 | rs34518860 | Intergenic | 0.87 | 4.99 _X_ 10^-08^ |  |  |
| chr6:32637266 | rs9272416 | Upstream | 0.91 | 9.25 _X_ 10^-09^ | *HLA-DQA1* | *-* |
| chr6:32639231 | rs9272553 | Intron | 0.92 | 1.48 _X_ 10^-08^ |  |  |
| chr6:32640052 | rs9272613 | Intron | 0.92 | 2.04 _X_ 10^-08^ |  |  |
| chr6:32640237 | rs9272625 | Intron | 0.92 | 2.84 _X_ 10^-08^ |  | *-* |
| chr6:32641328 | rs1129740 | Missense | 0.92 | 2.24 _X_ 10^-08^ |  |  |
| chr6:32641349 | rs1071630 | Missense | 0.92 | 2.48 _X_ 10^-08^ |  |  |
| chr6:32642282 | rs34843907 | Intron | 0.92 | 2.25 _X_ 10^-08^ |  |  |
| chr6:32646047 | rs9273226 | Intron | 0.92 | 1.39 _X_ 10^-08^ |  |  |
| chr6:32646972 | rs17612489 | Intron | 0.92 | 1.49 _X_ 10^-08^ |  |  |
| chr6:32647106 | rs17612510 | Intron | 0.92 | 2.40 _X_ 10^-08^ |  |  |
| chr6:32647750 | rs17843579 | Intron | 0.92 | 3.25 _X_ 10^-08^ | *HLA-DQA1* | *HLA-DRB1, HLA-DQA1, HLA-DQB1, HLA-DQA2* |
| chr6:32652567 | rs17612788 | Intron | 0.92 | 2.81 _X_ 10^-08^ |  |  |
| chr6:32652582 | rs17612802 | Intron | 0.92 | 1.94 _X_ 10^-08^ |  |  |
| chr6:32652845 | rs17612858 | Intron | 0.92 | 1.91 _X_ 10^-08^ |  |  |
| chr6:32653166 | rs17612921 | Intron | 0.92 | 2.09 _X_ 10^-08^ |  |  |
| chr6:32655325 | rs9273322 | Downstream | 0.92 | 1.64 _X_ 10^-08^ |  |  |
| chr6:32655442 | rs9273326 | Intergenic | 0.92 | 1.84 _X_ 10^-08^ | *HLA-DQA1-HLA-DQB1* |  |
| chr6:32655465 | rs9273329 | Intergenic | 0.92 | 1.98 _X_ 10^-08^ |  |  |
| chr6:32655523 | rs9273330 | Intergenic | 0.92 | 2.44 _X_ 10^-08^ |  |  |
| chr6:32656954 | rs74824383 | Intergenic | 0.92 | 1.90 _X_ 10^-08^ |  |  |
| chr6:32657081 | rs72852269 | Intergenic | 0.92 | 2.04 _X_ 10^-08^ |  |  |
| chr6:32659544 | rs9273416 | 3’UTR | 0.92 | 2.05 _X_ 10^-08^ | *HLA-DQB1* |  |
| chr6:32659657 | rs9273426 | 3’UTR | 0.92 | 1.95 _X_ 10^-08^ |  |  |
| chr6:32660129 | rs1063349 | 3’UTR | 0.92 | 3.00 _X_ 10^-08^ |  |  |
| chr6:32661360 | rs1140343 | Missense | 0.91 | 1.07 _X_ 10^-08^ |  |  |
| chr6:32666737 | rs9274529 | Intergenic | 0.92 | 4.79 _X_ 10^-08^ | *HLA-DQB1-MTCO3P1* |  |
| chr6:32667977 | rs3828789 | Intergenic | 0.92 | 1.72 _X_ 10^-08^ |  |  |
| chr6:32668032 | rs3828790 | Intergenic | 0.92 | 2.36 _X_ 10^-08^ |  |  |
| chr9:6209697 | rs992969 | Intergenic | 1.12 | 2.83 _X_ 10^-10^ | *GTF3AP1-IL33* | *IL33* |
| chr11:76588605 | rs11236797 | Regulatory | 1.10 | 4.24 _X_ 10^-10^ | *LINC02757-EMSY* | *LRRC32* |
| chr15:67150258 | rs17293632 | Intron | 1.12 | 1.64 _X_ 10^-08^ | *SMAD3* | *SMAD3, IQCH, AAGAB* |
| chr15:67157967 | rs17228058 | Intron | 1.12 | 1.75 _X_ 10^-08^ |  |  |
| chr17:39813896 | rs9909593 | Intron | 0.91 | 5.80 _X_ 10^-09^ | *IKZF3* | *IKZF3, GRB7, MIEN1, GSDMB* |
| chr17:39820216 | rs9303277 | Intron | 0.92 | 1.38 _X_ 10^-08^ |  |  |
| chr17:39905943 | rs2305480 | Missense | 0.92 | 3.95 _X_ 10^-08^ | *GSDMB* | *GSDMA, ORMDL3, GSDMB, LRRC3C, IKZF3, PSMD3* |
| chr17:39906723 | rs11078926 | Intron | 0.92 | 4.07 _X_ 10^-08^ |  |  |

BP = base-pair; SNP = single nucleotide polymorphism; OR = odds ratios; eQTL = expression quantitative trait locus

**Supplementary Table S2**: Genome-wide significant loci and lung eQTL candidate genes identified in the meta-analysis GWAS of mild asthma

| **BP** | **SNP** | **Location** | **OR** | **P** | **I^2^** | **Genes** | **Lung eQTL** |
| --- | --- | --- | --- | --- | --- | --- | --- |
| chr2:102275879 |  | Intergenic | 0.87 | 3.31E-08 | 50.6 | *CFAP144P2-IL1RL1* | *IL1RL1, IL18R1, IL1RL2* |
| chr2:102319514 | rs11693204 | Intron | 1.11 | 4.07 _X_ 10^-08^ | 0.0 | *IL1RL1* | *IL1RL1, IL18R1, IL18RAP* |
| chr2:102319699 | rs72823641 | Intron | 0.87 | 4.25 _X_ 10^-08^ | 22.4 |  |  |
| chr2:102332701 | rs12479210 | Intron | 1.10 | 4.40 _X_ 10^-08^ | 50.3 |  |  |
| chr2:102334362 | rs13019081 | Intron | 1.10 | 2.27 _X_ 10^-08^ | 45.5 |  |  |
| chr2:102337157 | rs3771180 | Intron | 0.88 | 2.09 _X_ 10^-09^ | 59.4 |  |  |
| chr2:102338622 | rs13408661 | Intron | 0.88 | 1.80 _X_ 10^-09^ | 59.1 |  |  |
| chr2:102339223 | rs873022 | Intron | 1.11 | 3.63 _X_ 10^-08^ | 0.0 |  |  |
| chr2:102339400 | rs3771177 | Intron | 1.11 | 4.07 _X_ 10^-08^ | 0.0 |  |  |
| chr2:102341072 | rs3732129 | Intron | 1.11 | 4.16 _X_ 10^-08^ | 0.0 |  |  |
| chr2:102341256 | rs1420101 | Intron | 1.10 | 5.45 _X_ 10^-09^ | 0.0 |  |  |
| chr2:102343547 | rs12905 | 3’UTR | 1.11 | 1.27 _X_ 10^-08^ | 0.0 |  |  |
| chr2:102343750 | rs3771175 | 3’UTR | 0.88 | 2.02 _X_ 10^-09^ | 60.3 |  |  |
| chr2:102343821 | rs3821204 | Intron | 1.11 | 3.91 _X_ 10^-08^ | 0.0 |  |  |
| chr2:102348401 | rs55927292 | Intron | 1.11 | 2.59 _X_ 10^-09^ | 0.0 |  |  |
| chr2:102348872 | rs17027006 | Intron | 1.11 | 2.85 _X_ 10^-08^ | 0.0 |  |  |
| chr2:102349411 | rs12469506 | Intron | 1.11 | 3.64 _X_ 10^-08^ | 0.0 |  |  |
| chr2:102350089 | rs10197862 | Intron | 0.88 | 2.95 _X_ 10^-09^ | 56.7 |  |  |
| chr2:102353347 | rs67723747 | Intergenic | 1.11 | 1.84 _X_ 10^-08^ | 0.0 | *IL1RL1-IL18R1* |  |
| chr2:102354705 | rs56386507 | Intergenic | 1.11 | 1.91 _X_ 10^-08^ | 0.0 |  |  |
| chr2:102369352 | rs3771172 | Intron | 1.11 | 2.74 _X_ 10^-08^ | 0.0 | *IL18R1* |  |
| chr2:102369490 | rs3771171 | Intron | 1.11 | 3.10 _X_ 10^-08^ | 0.0 |  |  |
| chr2:102369694 | rs2160202 | Intron | 1.11 | 3.14 _X_ 10^-08^ | 0.0 |  |  |
| chr2:102375619 | rs2270298 | Intron | 1.11 | 1.04 _X_ 10^-08^ | 0.0 |  |  |
| chr2:102378424 | rs17027037 | Intron | 1.11 | 8.75 _X_ 10^-09^ | 0.0 |  |  |
| chr2:102380345 | rs11683700 | Intron | 1.11 | 1.41 _X_ 10^-08^ | 0.0 |  |  |
| chr2:102380412 | rs3821203 | Intron | 1.11 | 1.04 _X_ 10^-08^ | 0.0 |  |  |
| chr2:102380714 | rs3771162 | Intron | 1.11 | 3.07 _X_ 10^-08^ | 0.0 |  |  |
| chr2:102381273 | rs11465633 | Intron | 1.11 | 1.31 _X_ 10^-08^ | 0.0 |  |  |
| chr2:102382852 | rs56258475 | Intron | 1.11 | 3.21 _X_ 10^-08^ | 36.9 |  |  |
| chr2:102384942 | rs1035130 | Intron | 1.11 | 1.74 _X_ 10^-08^ | 0.0 |  |  |
| chr2:102386805 | rs2241116 | Intron | 1.12 | 1.63 _X_ 10^-08^ | 0.0 |  |  |
| chr2:102389927 | rs4851570 | Intron | 1.11 | 1.26 _X_ 10^-08^ | 0.0 |  |  |
| chr2:102391232 | rs1680552504 | Intron | 1.11 | 1.66 _X_ 10^-08^ | 0.0 |  |  |
| chr2:102392509 | rs66919607 | Intron | 1.11 | 2.13 _X_ 10^-08^ | 0.0 |  |  |
| chr2:102394070 | rs2287035 | Intron | 1.11 | 2.46 _X_ 10^-08^ | 0.0 |  |  |
| chr2:102394128 | rs2287034 | Intron | 1.11 | 3.25 _X_ 10^-08^ | 4.2 |  |  |
| chr2:102396214 | rs17027071 | Intron | 1.11 | 1.80 _X_ 10^-08^ | 0.0 |  |  |
| chr2:102397842 | rs1135354 | 3’UTR | 1.11 | 2.08 _X_ 10^-08^ | 0.0 |  |  |
| chr2:102399458 | rs17027087 | Intergenic | 1.11 | 1.86 _X_ 10^-08^ | 0.0 | *IL18R1-SDR42E1P5* |  |
| chr2:102401617 | rs3732123 | Intergenic | 1.11 | 2.08 _X_ 10^-08^ | 0.0 |  |  |
| chr2:102403322 | rs55742125 | Intergenic | 1.11 | 2.62 _X_ 10^-08^ | 0.0 |  |  |
| chr2:102407871 | rs55883125 | Intergenic | 1.11 | 1.30 _X_ 10^-08^ | 0.0 |  |  |
| chr2:102408278 | rs4851006 | Intergenic | 1.11 | 2.04 _X_ 10^-08^ | 0.0 |  |  |
| chr2:102420217 | rs3771156 | Intron | 1.11 | 2.91 _X_ 10^-08^ | 0.0 | *IL18RAP* | *IL18RAP* |
| chr2:102431342 | rs66566526 | Intron | 1.11 | 2.60 _X_ 10^-08^ | 35.4 |  |  |
| chr2:102431697 | rs56331791 | Intron | 1.11 | 4.52 _X_ 10^-08^ | 47.2 |  |  |
| chr2:102434402 |  |  | 1.11 | 1.06 _X_ 10^-08^ | 0.4 |  |  |
| chr2:102434684 | rs11681718 | Intron | 1.11 | 2.94 _X_ 10^-08^ | 18.7 |  |  |
| chr2:102435098 | rs4851582 | Intron | 1.11 | 3.87 _X_ 10^-09^ | 0.0 |  |  |
| chr2:102436842 | rs58883541 | Intron | 1.11 | 1.23 _X_ 10^-09^ | 0.0 |  |  |
| chr2:102438960 | rs17027166 | Intron | 1.11 | 1.28 _X_ 10^-09^ | 0.0 |  |  |
| chr2:102439201 | rs55645612 | Intron | 1.11 | 1.35 _X_ 10^-09^ | 0.0 |  |  |
| chr2:102439636 | rs56166614 | Intron | 1.11 | 1.35 _X_ 10^-09^ | 0.0 |  |  |
| chr2:102440074 | rs10490204 | Intron | 1.11 | 2.07 _X_ 10^-08^ | 25.5 |  |  |
| chr2:102440699 | rs17027179 | Intron | 1.11 | 1.10 _X_ 10^-09^ | 0.0 |  |  |
| chr2:102459966 | rs11690532 | Intergenic | 1.13 | 1.84 _X_ 10^-08^ | 0.0 | *IL18RAP-SLC9A4* |  |
| chr2:102473668 | rs17027255 | 5’UTR | 1.11 | 9.22 _X_ 10^-10^ | 0.0 | *SLC9A4* |  |
| chr2:102475081 | rs17027258 | Intron | 1.12 | 7.62 _X_ 10^-10^ | 0.0 |  |  |
| chr6:32616804 | rs73730372 | Intergenic | 0.87 | 3.61 _X_ 10^-09^ | 29.5 | *HLA-DRB1-HLA-DQA1* | *HLA-DQB1, HLA-DQA1, HLA-DQB1-AS1, HLA-DRB5, HLA-DRB1* |
| chr6:32617294 | rs113243185 | Intergenic | 0.87 | 2.84 _X_ 10^-09^ | 17.9 |  |  |
| chr6:32622991 | rs34831921 | Intergenic | 0.87 | 1.49 _X_ 10^-08^ | 0.0 |  |  |
| chr6:32626326 | rs34518860 | Intergenic | 0.87 | 1.82 _X_ 10^-08^ | 0.0 |  |  |
| chr6:32631560 | rs34061722 | Intergenic | 0.90 | 7.75 _X_ 10^-09^ | 34.1 |  |  |
| chr6:32634653 | rs17211510 | Intergenic | 0.90 | 1.49 _X_ 10^-08^ | 41.8 |  |  |
| chr6:32635578 | rs28407322 | Upstream | 0.90 | 1.27 _X_ 10^-08^ | 37.7 | *HLA-DQA1-AS1* |  |
| chr6:32635711 | rs28675927 | Upstream | 0.90 | 7.93 _X_ 10^-09^ | 41.0 |  |  |
| chr6:32637266 | rs9272416 | Upstream | 0.91 | 1.60 _X_ 10^-09^ | 35.0 |  |  |
| chr6:32639231 | rs9272553 | Intron | 0.91 | 2.38 _X_ 10^-09^ | 29.2 |  |  |
| chr6:32640052 | rs9272613 | Intron | 0.91 | 3.43 _X_ 10^-09^ | 42.4 |  |  |
| chr6:32640237 | rs9272625 | Intron | 0.91 | 4.36 _X_ 10^-09^ | 43.8 |  |  |
| chr6:32641328 | rs1129740 | Missense | 0.91 | 3.54 _X_ 10^-09^ | 42.4 |  |  |
| chr6:32641349 | rs1071630 | Missense | 0.91 | 3.85 _X_ 10^-09^ | 43.5 | *HLA-DQA1* |  |
| chr6:32642282 | rs34843907 | Intron | 0.91 | 3.76 _X_ 10^-09^ | 41.8 |  |  |
| chr6:32646047 | rs9273226 | Intron | 0.91 | 2.38 _X_ 10^-09^ | 41.2 |  |  |
| chr6:32646303 | rs9273242 | Intron | 0.91 | 2.82 _X_ 10^-08^ | 37.5 |  |  |
| chr6:32646972 | rs17612489 | Intron | 0.91 | 2.21 _X_ 10^-09^ | 41.8 |  |  |
| chr6:32647106 | rs17612510 | Intron | 0.91 | 4.17 _X_ 10^-09^ | 41.6 |  |  |
| chr6:32647693 | rs17612583 | Intron | 0.91 | 1.30 _X_ 10^-08^ | 47.1 |  |  |
| chr6:32647750 | rs17843579 | Intron | 0.91 | 7.57 _X_ 10^-09^ | 50.2 |  |  |
| chr6:32652567 | rs17612788 | Intron | 0.91 | 4.25 _X_ 10^-09^ | 39.8 |  |  |
| chr6:32652582 | rs17612802 | Intron | 0.91 | 2.93 _X_ 10^-09^ | 42.0 |  |  |
| chr6:32652845 | rs17612858 | Intron | 0.91 | 3.59 _X_ 10^-09^ | 44.6 |  |  |
| chr6:32653166 | rs17612921 | Intron | 0.91 | 3.49 _X_ 10^-09^ | 40.8 |  |  |
| chr6:32655325 | rs9273322 | Intergenic | 0.91 | 2.64 _X_ 10^-09^ | 41.9 | *HLA-DQA1-* *HLA-DQB1* |  |
| chr6:32655442 | rs9273326 | Intergenic | 0.91 | 3.02 _X_ 10^-09^ | 44.5 |  |  |
| chr6:32655465 | rs9273329 | Intergenic | 0.91 | 3.20 _X_ 10^-09^ | 42.6 |  |  |
| chr6:32655523 | rs9273330 | Intergenic | 0.91 | 4.08 _X_ 10^-09^ | 42.2 |  |  |
| chr6:32656954 | rs74824383 | Intergenic | 0.91 | 3.25 _X_ 10^-09^ | 41.3 |  |  |
| chr6:32657081 | rs72852269 | Intergenic | 0.91 | 3.40 _X_ 10^-09^ | 44.4 |  |  |
| chr6:32659544 | rs9273416 | 3’UTR | 0.91 | 3.20 _X_ 10^-09^ | 39.2 | *HLA-DQB1* |  |
| chr6:32659657 | rs9273426 | 3’UTR | 0.91 | 3.31 _X_ 10^-09^ | 38.8 |  |  |
| chr6:32660129 | rs1063349 | 3’UTR | 0.91 | 4.70 _X_ 10^-09^ | 45.2 |  |  |
| chr6:32661360 | rs1140343 | Missense | 0.91 | 2.14 _X_ 10^-09^ | 42.0 |  |  |
| chr6:32666737 | rs9274529 | Intergenic | 0.91 | 7.64 _X_ 10^-09^ | 47.6 | *HLA-DQB1-MTCO3P1* |  |
| chr6:32667977 | rs3828789 | Intergenic | 0.91 | 2.62 _X_ 10^-09^ | 33.8 |  |  |
| chr6:32668032 | rs3828790 | Intergenic | 0.91 | 3.53 _X_ 10^-09^ | 41.4 |  |  |
| chr9:6193455 | rs2381416 | Intergenic | 1.11 | 2.22 _X_ 10^-09^ | 0.0 | *GTF3AP1-RANBP6* | *IL33* |
| chr9:6209697 | rs992969 | Intergenic | 1.11 | 5.53 _X_ 10^-09^ | 0.0 | *GTF3AP1-IL33* |  |
| chr9:6213387 | rs928413 | Intergenic | 1.11 | 1.25 _X_ 10^-09^ | 0.0 |  |  |
| chr11:76588605 | rs11236797 | Intergenic | 1.10 | 3.33 _X_ 10^-10^ | 9.9 | *LINC02757, EMSY* | *LRRC32* |
| chr15:67150258 | rs17293632 | Intron | 1.12 | 6.21 _X_ 10^-09^ | 5.3 | *SMAD3* | *SMAD3* |
| chr15:67157967 | rs17228058 | Intron | 1.12 | 6.63 _X_ 10^-09^ | 0.0 |  | *SMAD3, IQCH, AAGAB* |
| chr17:39813896 | rs9909593 | Intron | 0.92 | 4.85 _X_ 10^-08^ | 19.1 | *IKZF3* | *IKZF3, GRB7, MIEN1* |

BP = base-pair; SNP = single nucleotide polymorphism; OR = odds ratios; eQTL = expression quantitative trait locus

**Supplementary Table S3**: Genome-wide significant loci and lung eQTL candidate genes identified in the pooled GWAS of moderate-to-severe asthma

| **Position** | **SNP** |  | **OR** | **P** | **Gene** | **Lung eQTL** |
| --- | --- | --- | --- | --- | --- | --- |
| chr2:102316102 | rs950880 | Intron | 1.13 | 4.13 _X_ 10^-09^ | *IL1RL1* | *IL1RL1, IL18R1, IL18RAP* |
| chr2:102332701 | rs12479210 | Intron | 1.13 | 2.75 _X_ 10^-09^ |  |  |
| chr2:102334362 | rs13019081 | Intron | 1.14 | 1.77 _X_ 10^-09^ |  |  |
| chr2:102341256 | rs1420101 | Intron | 1.13 | 1.95 _X_ 10^-09^ |  |  |
| chr5:111116386 | rs6884870 | Intron | 1.14 | 1.61 _X_ 10^-09^ | *WDR36* | *WDR36, TSLP, CAMK4* |
| chr5:111117246 | rs77793850 | Intron | 1.14 | 3.15 _X_ 10^-09^ |  |  |
| chr5:111117378 | rs17624321 | Intron | 1.13 | 3.05 _X_ 10^-08^ |  |  |
| chr5:111121460 | rs17624673 | Intron | 1.13 | 4.13 _X_ 10^-08^ |  |  |
| chr5:111128310 | rs1043828 | 3’UTR | 1.14 | 4.09 _X_ 10^-09^ |  |  |
| chr5:111131801 | rs1438673 | Intergenic | 1.14 | 8.20 _X_ 10^-11^ | *WDR36-RPS3AP21* |  |
| chr5:111133279 | rs34962275 | Intergenic | 1.13 | 4.72 _X_ 10^-09^ |  |  |
| chr6:137432867 | rs147917037 | Intron | 1.77 | 1.17 _X_ 10^-08^ | *LOC102723633* |  |
| chr9:6209697 | rs992969 | Intergenic | 1.17 | 6.69 _X_ 10^-13^ | *GTF3AP1-IL33* | *IL33* |
| chr9:95513823 | rs28485705 | Intron | 0.86 | 4.08 _X_ 10^-08^ | *PTCH1* | *PTCH1, FANCC* |
| chr17:39820216 | rs9303277 | Intron | 0.89 | 4.05 _X_ 10^-09^ | *IKZF3* | *GSDMB, MIEN1, IKZF3, GRB7* |

BP = base-pair; SNP = single nucleotide polymorphism; OR = odds ratios; eQTL = expression quantitative trait locus

**Supplementary Table S4**: Genome-wide significant loci and lung eQTL candidate genes identified in the meta-analysis GWAS of moderate-to-severe asthma

| **Position** | **SNP** |  | **OR** | **P** | **I^2^** | **Gene** | **Lung eQTL** |
| --- | --- | --- | --- | --- | --- | --- | --- |
| chr2:102316102 | rs950880 | Intron | 1.13 | 8.96 _X_ 10^-09^ | 36.9 | *IL1RL1* | *IL1RL1, IL18R1, IL18RAP* |
| chr2:102332701 | rs12479210 | Intron | 1.14 | 5.20 _X_ 10^-09^ | 41.3 |  |  |
| chr2:102334362 | rs13019081 | Intron | 1.14 | 3.16 _X_ 10^-09^ | 46.5 |  |  |
| chr2:102341256 | rs1420101 | Intron | 1.13 | 4.05 _X_ 10^-09^ | 18.6 |  |  |
| chr5:111116386 | rs6884870 | Intron | 1.13 | 2.77 _X_ 10^-08^ | 0 | *WDR36* | *WDR36, CAMK4, TSLP* |
| chr5:111117246 | rs77793850 | Intron | 1.14 | 4.64 _X_ 10^-08^ | 0 |  |  |
| chr5:111128310 | rs1043828 | 3’UTR | 1.13 | 4.09 _X_ 10^-08^ | 0 |  |  |
| chr5:111131801 | rs1438673 | Intergenic | 1.14 | 6.65 _X_ 10^-10^ | 0 | *WDR36-RPS3AP21* |  |
| chr6:32618459 | rs11751024 | Intergenic | 0.89 | 3.25 _X_ 10^-08^ | 80.9 | *HLA-DRB1-HLA-DQA1* | *HLA-DRB1, HLA-DQA1, HLA-DRB5, HLA-DQB1, HLA-DQB1-AS1* |
| chr6:32625019 | rs28383322 | Intergenic | 0.87 | 3.23 _X_ 10^-08^ | 0 |  |  |
| chr9:6193455 | rs2381416 | Intergenic | 1.16 | 3.00 _X_ 10^-12^ | 19.3 | *GTF3AP1-RANBP6* | *IL33* |
| chr9:6209697 | rs992969 | Intergenic | 1.16 | 2.55 _X_ 10^-11^ | 5.0 | *GTF3AP1-IL33* |  |
| chr9:6213387 | rs928413 | Intergenic | 1.16 | 9.10 _X_ 10^-12^ | 3.8 | *GTF3AP1-IL33* |  |
| chr11:76588605 | rs11236797 | Regulatory | 1.12 | 4.47 _X_ 10^-08^ | 53.9 | *LINC02757-EMSY* | *LRRC32* |

BP = base-pair; SNP = single nucleotide polymorphism; OR = odds ratios; eQTL = expression quantitative trait locus

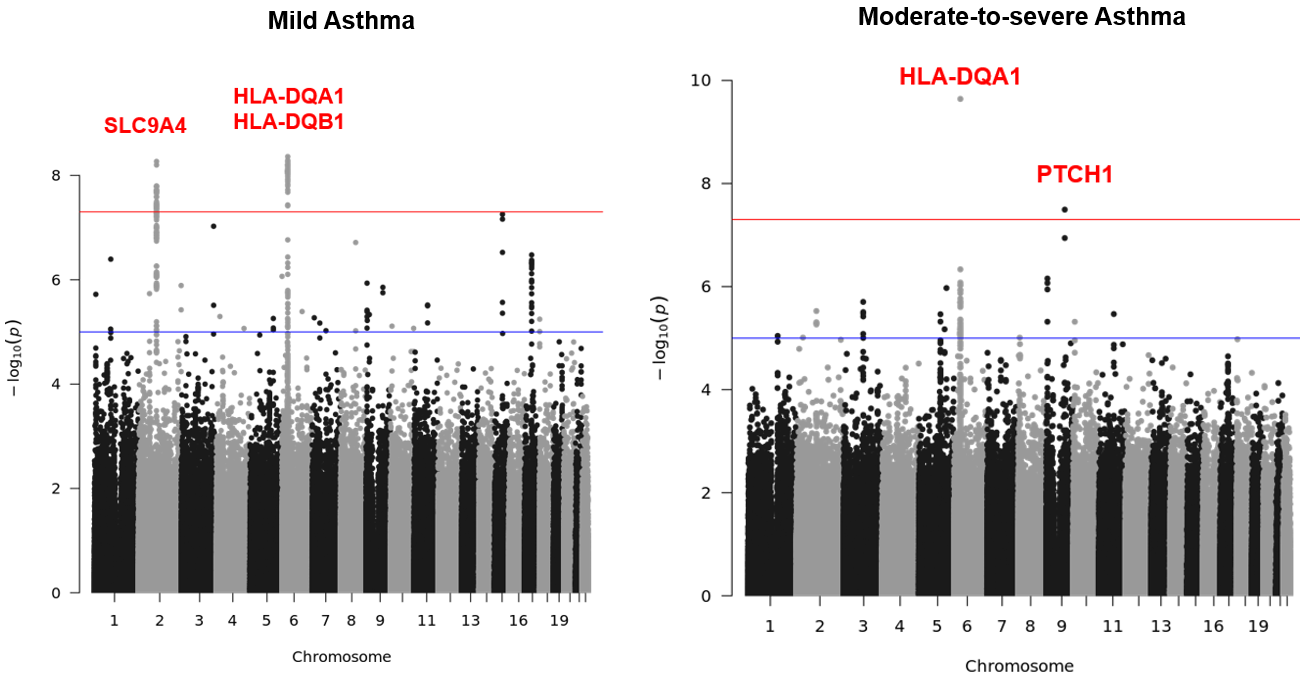

**Supplementary Figure S1**. Manhattan plots of the European ancestry-specific GWAS for mild and moderate-to-severe asthma. The red horizontal line indicates the genome-wide significance threshold (*P* < 5 × 10^-8^). The blue horizontal line indicates the suggestive significance threshold (*P* < 1 × 10^-5^). The *SLC9A4* and *HLA-DQA1/HLA-DQB1* loci reached genome-wide significance in mild asthma, whereas the HLA-DQA1 and PTCH1 loci reached genome-wide significance in moderate-to-severe asthma.

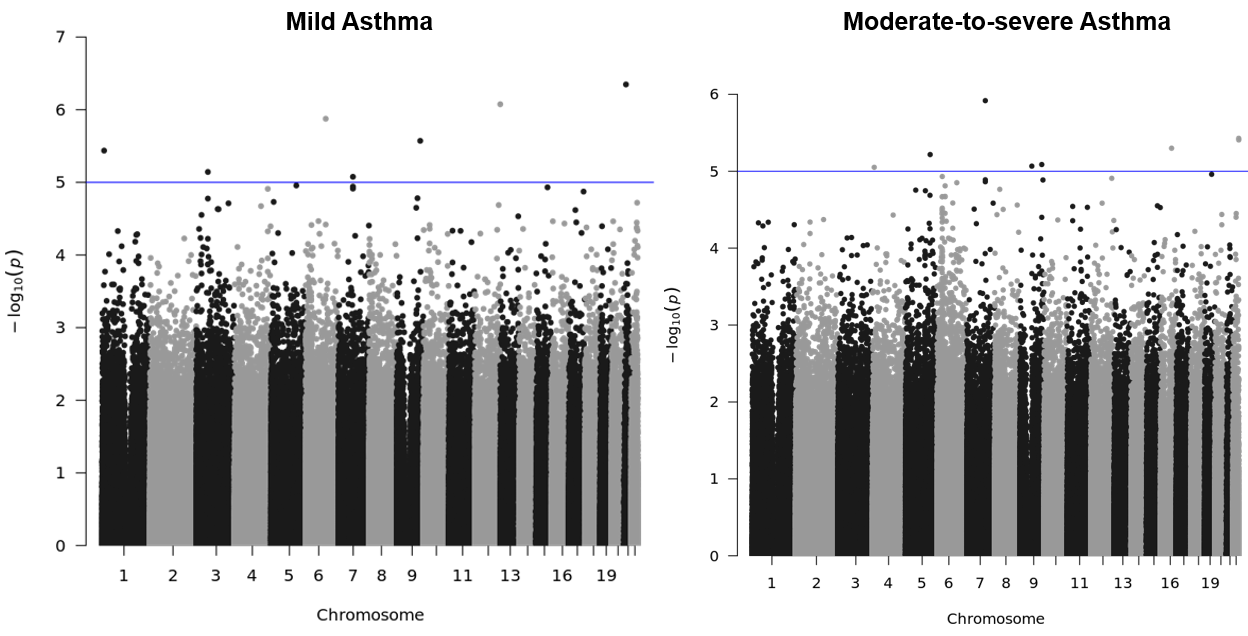

**Supplementary Figure S2**. Manhattan plots of the African ancestry-specific GWAS for mild and moderate-to-severe asthma. No locus reached genome-wide significance at *P* < 5 × 10^-8^. The blue horizontal line indicates the suggestive significance threshold (*P* < 1 × 10^-5^).

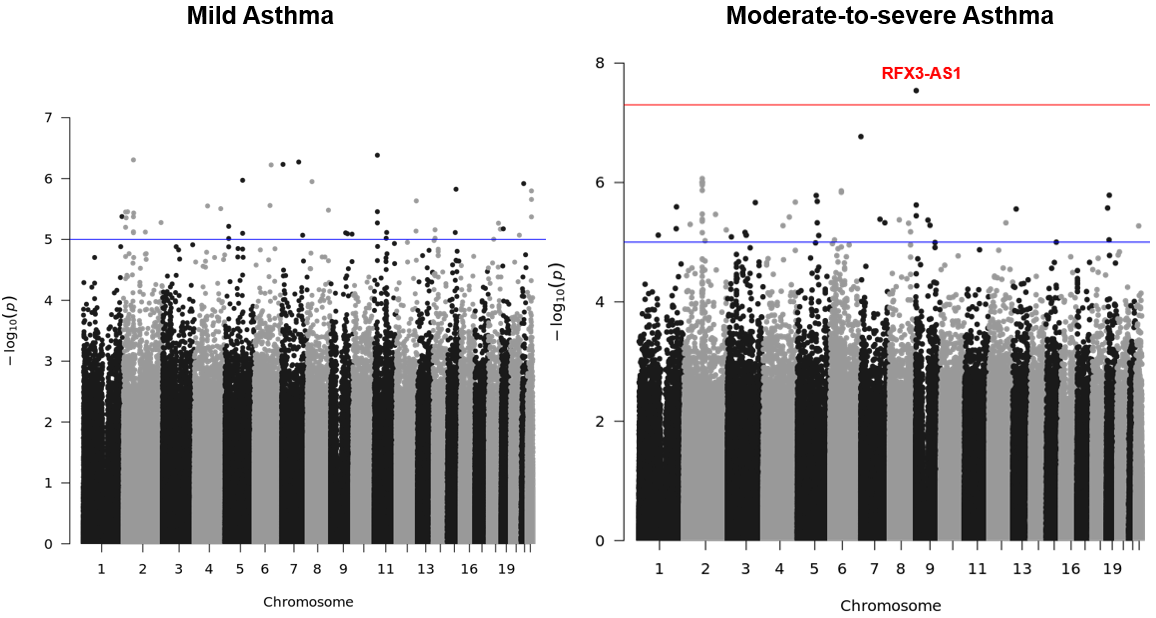

**Supplementary Figure S3**. Manhattan plots of Latino/Admixed ancestry-specific GWAS for mild and moderate-to-severe asthma. The red horizontal line indicates the genome-wide significance threshold (*P* < 5 × 10^-8^). The blue horizontal line indicates the suggestive significance threshold (*P* < 1 × 10^-5^). The *RFX3-AS1* locus reached genome-wide significance in the moderate-to-severe asthma analysis.
